# Accuracy, Utility and Applicability of the WHOOP Wearable Monitoring Device in Health, Wellness and Performance - a systematic review

**DOI:** 10.1101/2024.01.04.24300784

**Authors:** Riham Khodr, Latifah Kamal, Amir Minerbi, Gaurav Gupta

## Abstract

**Introduction:** The WHOOP wearable device is gaining popularity in clinical and performance applications with the ability to track sleep and heart rate parameters and provide feedback on recovery and strain. With the claims on potential benefits, a critical evaluation of the underlying scientific literature and the accuracy of these devices is imperative.

**Methods:** Authors systematically reviewed studies examining the accuracy and clinical applications of the WHOOP device.

**Results:** The WHOOP appears to have acceptable accuracy for two-stage sleep and heart rate metrics, but depending on the study, room for improvement for four-stage sleep and heart rate variability identification. There are numerous preliminary studies looking at the WHOOP’s ability to track and/or influence sleep and exercise behaviours at the cohort and/or population level. The impact of athletic performance and/or objective sleep is limited based on existing studies.

**Discussion:** The clinical application for the WHOOP, given the acceptable accuracy levels, continues to expand. Uses have included impact on sports performance, correlation with medical conditions (i.e. cognitive dysfunction), sleep and health behaviours in various populations. Limitations of existing accuracy trials include variable design and reporting metrics, while results from non-accuracy trials require further clinical validation for response rate and effect size.

**Conclusion:** The WHOOP wearable device has acceptable accuracy for sleep and cardiac variables to be used in clinical studies where a baseline can be established and, ideally, other clinical outcomes and gold standard tools can be employed.

## INTRODUCTION

Irrespective of the clinical or training setting, the optimal goal of monitoring physiological metrics is to optimize performance and wellness while preventing negative health outcomes (Kellmann, 2010), (Lundstrom, 2020). As technology evolves, so does the ability to provide non-invasive, portable and real-time feedback for sleep, heart variability (HRV) and activity levels (Plews et al., 2013, Prinsloo et al., 2014). The goal would be to use this data to inform decision-making to the benefit of the person tracking this information. For instance, high-performing groups (e.g., athletes, military, first responders) may benefit from automated feedback on physiological variables to direct activities and optimize preparedness and performance (Stone et al., 2020).

A variety of activity monitors are commercially available. Of these, the WHOOP wearable device (WHOOP Inc., Boston, MA) measures multiple variables and provides information on sleep, recovery and strain. Sleep quality (sleep disturbances and cycles), quantity (hours) and recovery (resting heart and rate heart rate variability) are used to provide the recovery score (i.e. ‘readiness to perform’). Strain uses continuous heart rate to measure energy expenditure and predict cardiovascular load during a given activity or throughout the day. (Lundstrom, 2020)

Although the WHOOP does not specifically measure energy metabolism, heart rate variability and sleep are sensitive to hemodynamic, endocrine, thermoregulatory, environmental, and psychological factors (A. Flatt et al., 2018; Lundstrom, 2020). In addition, training type, intensity and phase, baseline fitness, and body mass can also impact HRV and sleep metrics (Fortes et al., 2017); (Lundstrom, 2020) Despite more data being required, HRV and sleep appear to be correlated with reductions in athletic performance. (A. Flatt et al., 2018; A. A. Flatt & Howells, 2019; VanHeest et al., 2014; Woods et al., 2018, Woods et al., 2017, Reed et al., 2013, Sekiguchi et al., 2019, Bolin, 2019). Insufficient rest and sleep can impact cognitive and physical regeneration; therefore, identifying deficits could potentially impact outcomes (Dinges et al., 1997; Edwards & Waterhouse, 2009; Van Dongen et al., 2003; Vgontzas et al., 2004).

Therefore, using these tools to examine the autonomic system responsiveness to physiological stress could theoretically help examine the body’s ability to adapt to exercise stimulus and changes in health (C. R. Bellenger et al., 2016; Borresen & Lambert, 2008; Buchheit, 2014). While novel, the literature on the accuracy and clinical utility of these measures is still evolving. Studies on accuracy have looked at validation against gold standards, such as polysomnography (PSG) for sleep and electrocardiogram for heart rate variability (Berryhill et al., 2020). Other studies have looked at WHOOP use in performance, clinical prediction and treatment (Berryhill et al., 2020; Lundstrom, 2020).

While numerous limitations associated with “gold standard” measures of sleep and HRV (i.e. electrocardiograms, polysomnography, sleep surveys and diaries, actigraphy) exist, verification of commercially available activity trackers is still required (Depner et al., 2020; De Zambotti et al., 2019; Ibáñez et al., 2019; Kelly et al., 2012; Khosla et al., 2018; Stone et al., 2020). In this review, we look to systematically summarize the existing literature regarding the WHOOP wearable device concerning accuracy and applications.

## METHODS

A systematic literature search was conducted on Google, Google Scholar and PubMed by 3 authors (RK, LK and GG) using keywords WHOOP, activity trackers, sleep tracking, heart rate variability, performance, clinical applications, validation and accuracy. Search strategy included “WHOOP” OR “activity trackers” OR “accuracy” OR “validation” OR “clinical applications” OR “sleep tracking” OR “heart rate variability” OR “performance” OR “sleep tracking”.

Inclusion criteria were studies published in English, original research articles evaluating the accuracy of the WHOOP activity tracker, studies investigating the clinical applications or implications of using the WHOOP activity tracker in healthcare settings, studies conducted on human subjects of any age or health conditions and studies published from January 2000 to December 2023. Exclusion criteria were studies not written in English. reviews, meta-analyses, and opinion articles, studies not focusing on the WHOOP activity tracker, studies with insufficient data on accuracy or clinical applications and studies not conducted on human subjects.

We developed a standardized data extraction sheet to collect relevant information from each included study. This included fields for study characteristics (title, authors, publication year), study design, sample size, participant characteristics, WHOOP version/model, accuracy metrics, clinical outcomes, and any additional relevant information.

Given the number and heterogeneity of the studies, it was not possible to assess publication bias by means of any formal statistical tests.

A critical review was undertaken of these studies by the research team. Data was extracted from each article and prepared for a comparative analysis. Given the heterogeneity of the data, a meta-analysis of the data was not possible. Therefore, the analysis focused on a semi-quantitative and qualitative comparison looking at broad categories of accuracy and clinical/performance studies.

For accuracy studies, this review focused specifically on mean error measurements and not median error, limits of agreement, interquartile range or R^2^. Data was extracted from the publication and supplementary data. For clarification or corrections, multiple primary authors were contacted with no response. Therefore, adjustments to the presented data were made with consensus from the research team and, where possible, consistent with other data presented (Miller et al., 2020, Miller et al., 2021). Where manual and automatic detection were assessed separately, result ranges were displayed, and the absolute error was converted to percentage by dividing by the PSG measurements (Miller et al., 2021). Where only figures and no numbers were provided, calculations of mean absolute error were not possible (Grandner et al., 2023). The classification of bias for heart rate (HR) and HRV measures was done according to the following criteria proposed by Miller et al. 2022): 0.0–0.1, trivial; 0.1–0.3, small; 0.3–0.5, moderate; 0.5–0.7, large; 0.7–0.9, very large; 0.9–1.0, nearly perfect (C. Bellenger et al., 2021; Hopkins et al., 2009). Even with these guidelines, interpreting the sleep and HR/HRV outcomes was not easily possible from the available data in specific studies (Miller et al., 2022).

In addition, the values for bias did not correspond across all studies, nor did the reported analysis allow for standardization. Finally, commercial devices generally do not disclose their algorithms for detecting sleep states; therefore, light sleep was defined as the sum of N1 and N2, rapid eye movement (REM) and deep sleep (slow wave sleep) as N3 (Depner et al., 2020; Stone et al., 2020).

## RESULTS

50 abstracts were identified, and 11 relevant studies were selected for further review. From these 11 articles, 4 additional studies were found in the references, for a total of 15 studies included in this review. See Figure 1 for flowchart.

**Figure 1.**
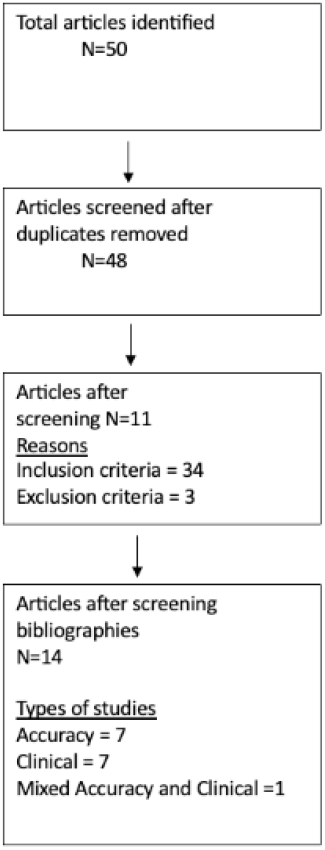
Article Selection Flowchart

The results have been divided into studies looking at WHOOP accuracy validation and performance/clinical application.

### Accuracy

Stone et al. showed a wide variation amongst the commercial devices tested, with error rates increasing for staging sleep (Stone et al., 2020). Miller et al. showed similar trends for sleep, wake and staging when comparing PSG to the WHOOP (Miller et al., 2020, 2022).

Bellenger et al. showed that WHOOP data required filtering to optimize ECG agreement, and correct interval identification has a more significant impact on HRV than HR calculations (C. Bellenger et al., 2021). Logarithmic HRV transformation also appears to reduce percent bias and residuals, which in part explains its use in monitoring training status (C. Bellenger et al., 2021). Inaccuracies in WHOOP-derived slow wave sleep (SWS)/deep sleep identification may impact HR and HRV measures modulated at this sleep stage (Burgess et al., 2004; Cabiddu et al., 2012; Gronfier et al., 1999; Somers et al., 1993). In one study, WHOOP-derived SWS episodes limits of agreement (LOA) for heart rate and HRV approached and/or exceeded the smallest worthwhile change and coefficient of variation, which may, in turn, impact the day-to-day variability in WHOOP-derived HR and HRV (C. Bellenger et al., 2021).

Miller et al. concluded that “comparisons of reliability based on intraclass correlations should be made across devices within the same study as there is no clear threshold at which a device can be considered valid” (Miller et al., 2021). Furthermore, both WHOOP manual and automatic two and four sleep categorization could provide a practical alternative to PSG and perform well against other commercially available devices (Miller et al., 2021). Grandner et al. showed moderate and comparable sleep specificity with commercially available activity trackers and that personalized algorithms that adapt to user data over time can improve device performance (Grandner et al., 2023).

See Tables 1 and 2 for more information regarding accuracy studies.

**Table 1:** Accuracy Studies Looking at Sleep Metrics and WHOOP Versus a Standard.

|  | N | Comparison | Generation | Total Sleep Time (TST) |  |  | Total Wake Time (TWT) |  |  | Light Sleep |  |  | Deep Sleep |  |  | REM |  |  |
| --- | --- | --- | --- | --- | --- | --- | --- | --- | --- | --- | --- | --- | --- | --- | --- | --- | --- | --- |
|  |  |  |  | MAPE (%) | Bias | Sensitivity (%) | MAPE (%) | Bias | Sensitivity (%) | MAPE (%) | Bias | Sensitivity (%) | MAPE (%) | Bias | Sensitivity (%) | MAPE (%) | Bias | Sensitivity(%) |
| (Stone et al., 2020) | 5 | HBSMS & various CAT | 2.0 | 8.8 | 0.27 |  | 60.3 |  | -0.59 | 13.5 | -0.17 |  | 46.2 | 0.15 |  | 57.8 | 0.10 |  |
| (Miller et al., 2020) | 12 | PSG | 2.0 | 2.3 | *22.5* | 95 | 15.1 | *22.5* | 51 | 2.1 | *34.6* | 62 | 4.4 | *20.1* | 68 | 17.6 | *34.1* | 70 |
| (Miller et al., 2021) | 6 | Actigraphy & PSG | 2.0 | 6.4-10.1 | -178.8 to 16.7 | 90-97 | 47.1-74.2 | -17.8 to 16.7 | 45-60 | 22.2-23.8 | -8.9 to 13.9 | 61-67 | 20.4-24.4 | -15.5 to -6.6 | 61-63 | 35.0 | 6.5-8.8 | 66 |
| (Grandner et al., 2023) | 36 | PSG, EEG headband, CAT | 3.0 | NC |  | 90-91 | NC |  |  | NC |  |  | NC |  |  | NC |  |  |
| (Berryhill et al., 2020) | 32 | PSG | 2.0 | *4.2* |  |  |  |  |  | NC |  |  | NC |  |  | *0.01* |  |  |
| (Miller et al., 2022) | 53 | PSG, ECG & CAT | 3.0 |  | -12.2 | 94 |  | 13.1 | 56 |  | -15.6 | 58 |  | -19.6 | 62 |  | 22.9 | 66 |

Mean Absolute Percentage Error (MAPE), Polysomnogram (PSG), Commercial Activity trackers (CAT), Home Based Sleep Monitoring Systems (HBSMS) , Electroencephalography (EEG), Not Calculable (NC). Electrocardiogram (ECG).
*\* NOTE: Data in red/italics could not be clarified*

**Table 2:** Accuracy Studies Looking at Cardiac Metrics and WHOOP Versus a Standard.

|  | N | Generation | Standard | Time Matched |  | Time Matched |  | Stage Matched |  |
| --- | --- | --- | --- | --- | --- | --- | --- | --- | --- |
|  |  |  |  | Bias HR - | Bias HR - LOA | Bias HRV - | Bias HR - LOA | HR/HRV Bias- | HR/HRV Bias- LOA |
| (C. Bellenger et al., 2021) | 6 | 2.0 | ECG/PSG | trivial | trivial | trivial | small | trivial | Moderate to large |
| (Berryhill et al., 2020) | 32 | 2.0 | PSG | *trivial* |  |  | *small* |  |  |
| (Miller et al., 2022) | 53 | 3.0 | PSG | -0.3 | 1.9 | -4.5 | 7.6 |  |  |

Polysomnogram (PSG), Electrocardiogram (ECG), Effect Size (ES), Commercial Activity trackers (CAT), Home Based Sleep Monitoring Systems (HBSMS)
*\* NOTE: Data in red/italics could not be clarified*

### Performance/Clinical Tracking

In studies looking at accuracy and clinical impact together, one study concluded that activity trackers can improve sleep quality and accurately measure sleep and cardiorespiratory variables (Berryhill et al., 2020). However, the impact of the WHOOP on athletic performance requires further investigation (Harms, 2018; Lundstrom, 2020).

There is emerging data suggesting HRV and time spent in slow wave sleep could correlate with cognitive function, thereby providing a non-invasive monitoring tool in pre-clinical cognitive impairment (Saif et al., 2019). Another study identified sleep patterns consistent with acute and chronic sleep deprivation amongst surgeons, declining post-call day 2 and recovering after post-call day 3 (Coleman et al., 2019). Additionally, poor sleep quality and quantity were seen in a group of orthopedic surgeons in another cohort study (Sochacki et al., 2018). Furthermore, one large-scale population monitoring showed a positive impact of physical distancing on sleep and exercise activity (Capodilupo & Miller, 2020), but a potential adverse association with mental health issues where sleep parameters are impaired (Czeisler et al., 2022).

See Table 3 for more information regarding the clinical and performance studies using the WHOOP.

**Table 3:**
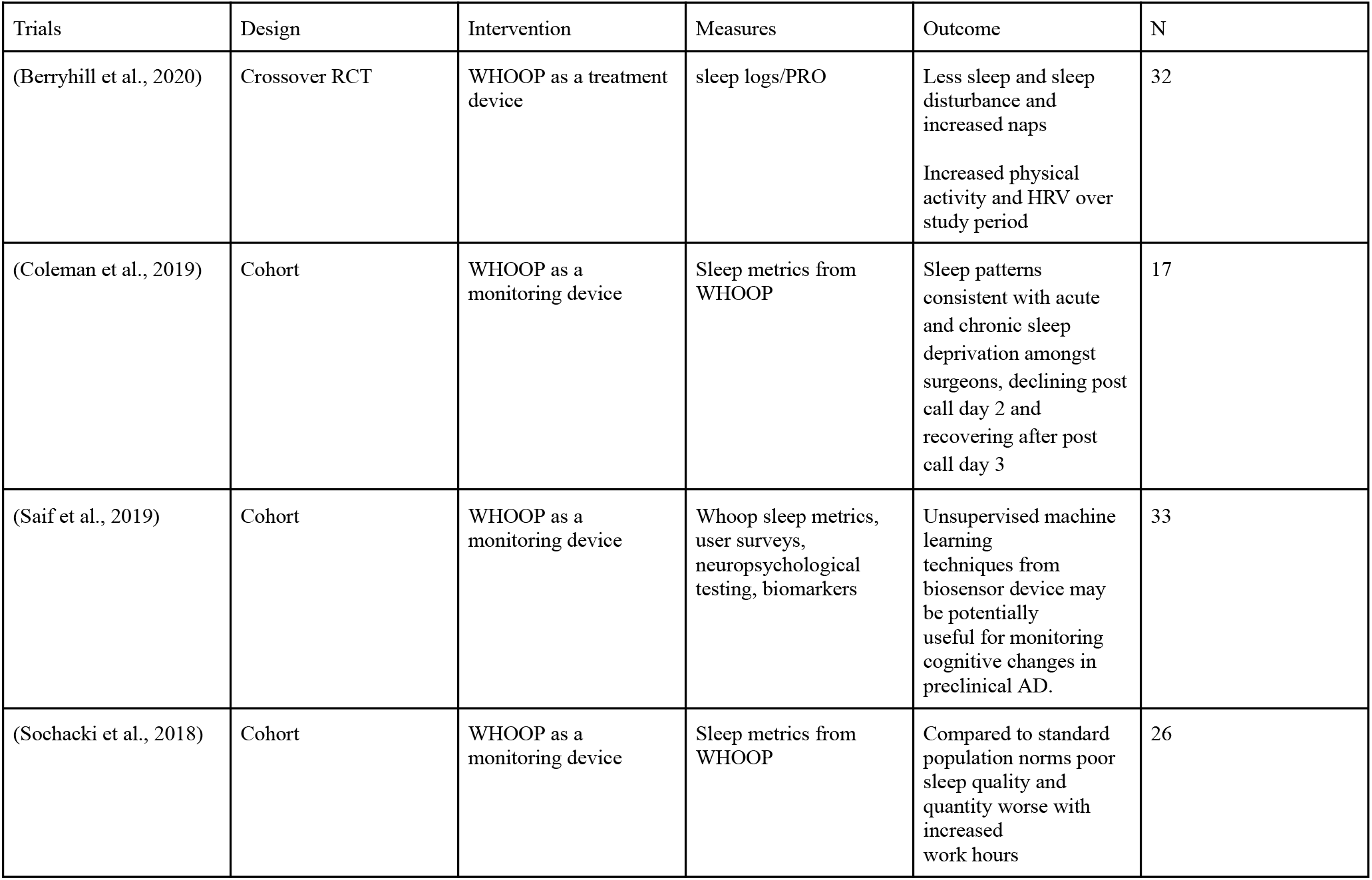
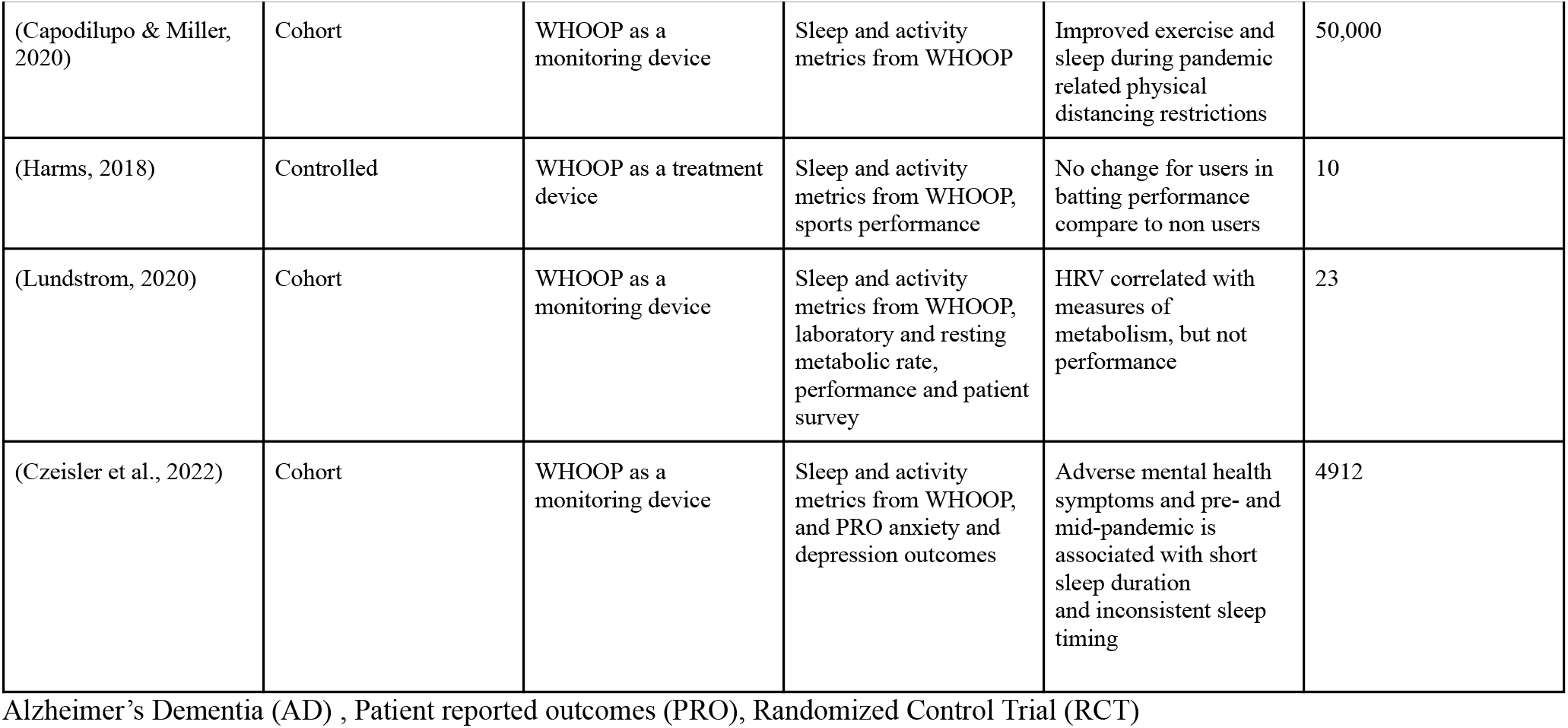
Clinical and Performance Studies looking at the application of WHOOP.

## DISCUSSION

In the setting of health and performance, using wireless and portable devices like the WHOOP to measure autonomic nervous system adaptability (HRV) and sleep quality affords numerous advantages compared to electrical-based cardiac measures and polysomnography (Kellmann, 2010; Plews et al., 2013; Prinsloo et al., 2014, D’Souza et al., 2014, Georgiou et al., 2018; Lai & Kim, 2015; Luczak et al., 2020; Luedtke & Duoos, 2015, Kellmann, 2010; Plews et al., 2013; Prinsloo et al., 2014, Georgiou et al., 2018; Lai & Kim, 2015; Luczak et al., 2020; Luedtke & Duoos, 2015).

The clinical application for the WHOOP, given the acceptable accuracy levels, continues to expand. Uses have included impact on sports performance, correlation with medical conditions (i.e. cognitive dysfunction), sleep and health behaviours in various populations. Limitations of existing accuracy trials include variability in study design and reporting metrics, while results from non-accuracy trials require further clinical validation of response rate and effect size.

### WHOOP Basics

The WHOOP uses plethysmography (PPG) to detect blood flow and heart rate changes between systole and diastole to calculate HRV (i.e. root mean square successive beat-to-beat interval differences) during slow wave sleep (Georgiou et al., 2018; Lai & Kim, 2015; Márquez & Molinero, 2013; Papageorgiou et al., 2018; Reed et al., 2013, D’Souza et al., 2014, Georgiou et al., 2018; Lai & Kim, 2015; Márquez & Molinero, 2013; Papageorgiou et al., 2018; Reed et al., 2013). Slow-wave sleep is theorized to play an integral role in physiological exercise recovery, therefore accurately measuring cardiac measures like HRV and heart rate could provide useful real-time feedback on the selection and monitoring of training activities (Gibbs et al., 2013; Papageorgiou et al., 2018; Scheid et al., 2009; Wade & Schneider, 1992, D’Souza et al., 2014, Georgiou et al., 2018; Lai & Kim, 2015; Márquez & Molinero, 2013; Papageorgiou et al., 2018; Reed et al., 2013, Gibbs et al., 2013; Papageorgiou et al., 2018; Scheid et al., 2009; Wade & Schneider, 1992). This hypothesis is supported by the correlation of certain sleep periods and growth hormone release, suggesting an optimization could improve performance and recovery (Shapiro et al., 1981).

### Sleep Tracking Limitations

Previous studies of activity trackers capable of identifying four-stage sleep data have generally yielded high sensitivity when an individual is asleep, low to moderate sensitivity for someone being awake, and varying sensitivity for various stages of sleep (De Zambotti et al., 2019; Miller et al., 2020; Shambroom et al., 2012). PPG-based technologies have historically been prone to errors, in part through motion artifacts and variability in skin complexion (Allen, 2007; Bent et al., 2020; Butler et al., 2016; Sañudo et al., 2019). Extracting data from commercial platforms can be problematic since multiple measures can be related, and standard deviations for each specific measure can influence the error measure (Stone et al., 2020).

In general, validation studies will also manually adjust activity tracker data in a research setting, meaning the autodetection accuracy may not be tested (de Zambotti et al., 2016, 2019; Kang et al., 2017; Maskevich et al., 2017; Meltzer et al., 2015; Miller et al., 2020). Even in the case of polysomnograms, technician-guided sleep staging can vary by up to 20%, which could influence the outcomes of studies on commercially available sleep and activity trackers (Collop, 2002). Therefore, the accuracy of sleep wearables in situations where manual adjustment of sleep times is performed by the user may vary. Finally, most accuracy validation studies have been done in healthy patients, with small sample sizes and variable nights of sleep, which may limit the generalizability of use (Miller et al., 2020).

### Heart Rate and Heart Rate Variability Limitations

For HR and HRV measures, there is likely a natural day-to-day variability of 3-13% related in part to timing and body position, as well as pulse travel to the periphery (Al Haddad et al., 2011; C. Bellenger et al., 2021; Chen et al., 2020; A. A. Flatt & Howells, 2019; Hopkins et al., 2009; Nakamura et al., 2020; Plews et al., 2013; Selvaraj et al., 2008). Therefore a filter is likely appropriate where bias and level of agreement are less than the smallest worthwhile change and coefficient of variability. However, where the bias and LOA reach or exceed the smallest worthwhile change/coefficient of variation, these values may need to be interpreted against a device’s own bias precision levels (Al Haddad et al., 2011; C. Bellenger et al., 2021; Chen et al., 2020; A. A. Flatt & Howells, 2019; Hopkins et al., 2009; Nakamura et al., 2020; Plews et al., 2013).

### Relevance Future Direction

Therefore, given the inaccuracies and variability, context could guide use. For commercial applications for the average person, the error rates are not expected to impact decisions around sleep and activity selection significantly. For research including baseline measures, a raw data feed, a gold standard where possible, other clinical variables, and devices with accelerometers to account for body movement, those using higher wavelength PPG could potentially reduce bias as the technologies and algorithms continue to evolve (Stone et al., 2020). In order to address the challenges with current research, some authors have recommended standard reporting that clinicians can easily understand and apply to clinic settings and monitor for over-focusing on sleep metrics in a way that creates adverse health effects such as insomnia (Khosla & Wickwire, 2020).

In “treatment” trials, the clinical utility and plausibility of outcomes should be examined closely. Even where the effect size appears to be small, how reduced overall sleep and increased nap time biologically correspond to patients’ reports of less sleep disturbance should be explored further in larger, participant-blinded trials (Berryhill et al., 2020). However, monitoring for sleep deprivation and/or cognitive decline in at-risk populations could be a strong value proposition for these tools, especially where outcomes are modifiable in a clinically relevant way (Coleman et al., 2019; Saif et al., 2019; Sochacki et al., 2018). Furthermore, based on current studies measuring population-level exercise and sleep data could be helpful from a public policy and planning standpoint (Capodilupo & Miller, 2020), but its impact on performance (e.g. sports, military, etc) needs to be clarified further (Harms, 2018; Lundstrom, 2020)

## CONCLUSIONS

The WHOOP wearable device has acceptable accuracy for sleep and cardiac variables to be used in clinical studies where a baseline can be established and ideally other clinical outcomes and gold standard tools can be employed. Further study with easy-to-understand standardized measures is required to understand the accuracy of newer devices and algorithms, as well as their utility in clinical decision-making and outcomes.

## Data Availability

All data produced in the present work are contained in the manuscript

